# Genetic susceptibility to schizophrenia through neuroinflammatory pathways is associated with retinal thinning: Findings from the UK-Biobank

**DOI:** 10.1101/2024.04.05.24305387

**Authors:** Finn Rabe, Lukasz Smigielski, Foivos Georgiadis, Nils Kallen, Wolfgang Omlor, Matthias Kirschner, Flurin Cathomas, Edna Grünblatt, Steven Silverstein, Brittany Blose, Daniel Barthelmes, Karen Schaal, Jose Rubio, Todd Lencz, Philipp Homan

**Affiliations:** Department of Adult Psychiatry and Psychotherapy, University of Zurich, Zurich, Switzerland; Department of Child and Adolescent Psychiatry and Psychotherapy, University Hospital of Psychiatry Zurich, University of Zurich, Zurich, Switzerland; Neuroscience Center Zurich, University of Zurich and ETH Zurich, Zurich, Switzerland; Zurich Center for Integrative Human Physiology, University of Zurich, Zurich, Switzerland; Department of Psychiatry, University of Rochester Medical Center, Rochester, New York, USA; Department of Ophthalmology, University of Rochester Medical Center, Rochester, New York, USA; Department of Neuroscience, University of Rochester Medical Center, Rochester, New York, USA; Center for Visual Science, University of Rochester, Rochester, New York, USA; Department of Ophthalmology, University Hospital Zurich, University of Zurich, Zurich, Switzerland; Department of Ophthalmology, Inselspital University Hospital Bern, Bern, Switzerland; Institute of Behavioral Science, Feinstein Institutes for Medical Research, Manhasset, NY, USA; Division of Psychiatry Research, Zucker Hillside Hospital, Northwell Health, New York, NY, USA; Department of Psychiatry, Zucker School of Medicine at Hofstra/Northwell, Hempstead, NY, USA

## Abstract

The human retina is part of the central nervous system and can be easily and non-invasively imaged with optical coherence tomography. While imaging the retina may provide insights on central nervous system-related disorders such as schizophrenia, a typical challenge are confounders often present in schizophrenia which may negatively impact retinal health. Here, we therefore aimed to investigate retinal changes in the context of common genetic variations conveying a risk of schizophrenia as measured by polygenic risk scores. We used population data from the UK Biobank, including White British and Irish individuals without diagnosed schizophrenia, and estimated a polygenic risk score for schizophrenia based on the newest genome-wide association study (PGC release 2022). We hypothesized that greater genetic susceptibility to schizophrenia is associated with retinal thinning, especially within the macula. To gain additional mechanistic insights, we conducted pathway-specific polygenic risk score associations analyses, focusing on gene pathways that are related to schizophrenia. Of 65484 individuals recruited, 48208 participants with available matching imaging-genetic data were included in the analysis of whom 22427 (53.48%) were female and 25781 (46.52%) were male. Our robust principal component regression results showed that polygenic risk scores for schizophrenia were associated with retinal thinning while controlling for confounding factors (b = −0.03, p = 0.007, pFWER = 0.01). Similarly, we found that polygenic risk for schizophrenia specific to neuroinflammation gene sets revealed significant associations with retinal thinning (b = −0.03, self-contained p = 0.041 (reflecting the level of association), competitive p = 0.05 (reflecting the level of enrichment)). These results go beyond previous studies suggesting a relationship between manifested schizophrenia and retinal phenotypes. They indicate that the retina is a mirror reflecting the genetic complexities of schizophrenia and that alterations observed in the retina of individuals with schizophrenia may be connected to an inherent genetic predisposition to neurodegenerative aspects of the condition. These associations also suggest the potential involvement of the neuroinflammatory pathway, with indications of genetic overlap with specific retinal phenotypes. The findings further indicate that this gene pathway in individuals with a high polygenic risk for schizophrenia could contribute through acute-phase proteins to structural changes in the retina.

## Introduction

The retina is a direct extension of the brain that allows for a non-invasive and real-time means of characterizing the neurovascular structure and function of the central nervous system.^1^ Recent studies using optical coherence tomography found retinal thinning in individuals with schizophrenia, suggesting that illness pathophysiology may be detected in the most distal and hence readily accessible part of the central nervous system.^2–8^ These studies have shown inner retinal atrophy, thinning of peripapillary retinal nerve fiber layers and macular ganglion cell and inner plexiform layers, as well as an enlarged cup-disc ratio. This was also confirmed in a large and well-characterized cohort,^9^ where individuals with schizophrenia showed an enlargement in cup disc ratio and a decrease in inner retinal thickness compared to healthy controls, beyond what could be attributed to the presence of hypertension and diabetes.^9^

Findings such as these underscore the retina’s relevance in understanding the neurobiological underpinnings of schizophrenia. However, an issue with many of these studies was the presence of potentially confounding factors such as the effects of antipsychotic medication, smoking, lifestyle factors, and disease-related changes, all which can negatively impact retinal health. Such confounders can in turn obscure the interpretation of findings, making it difficult to determine whether observed differences in retinal thickness are directly related to the pathophysiology of schizophrenia or are secondary to these confounding factors. In addition, retinal changes in these studies were associated with the disorder already being present, but the question remains whether these changes occurred before the onset of the disorder.

Intriguingly, retinal changes have been observed not only in patients but also in unaffected first-degree relatives, suggesting a link to genetic susceptibility to schizophrenia.^3^ Polygenic risk scores are an alternative to conventional heritability studies by allowing researchers to investigate the genetic underpinnings of retinal thickness changes in the context of schizophrenia risk, thus providing a potential understanding of the genetic contributions to these changes.^10^ They aggregate the impact of numerous genetic variants throughout the genome and account for a considerable portion of the variance in disease risk.^11,12^ The identification of shared genetic influences between retinal structures and schizophrenia^13,14^ further supports the hypothesis that retinal changes observed in schizophrenia could reflect underlying genetic susceptibilities. This convergence from optical coherence tomography studies and genetic research may help exploring how genetic predispositions contribute to the neurodevelopmental and neurodegenerative anomalies in schizophrenia^15^, including retinal alterations.

The present study aimed to understand whether higher polygenic risk for schizophrenia in healthy individuals is reflected in thinning of the retina. We focused on the macula, the area with the highest density of neurons in the retina (Figure 1). Different subfields of the macula may have different functions and may be affected differently by various diseases or conditions^16,17^ and may therefore be more informative when investigating polygenic associations. Furthermore, we explored gene set-specific polygenic risk scores for schizophrenia: biological pathways that are related to neurotransmitter regulation, inflammation and microvasculature, all of which may be altered in individuals with schizophrenia.^18–21^ Pathway-specific analyses focused on the cumulative genetic risk within specific biological pathways, which may offer insights into the heterogeneity of the disease and its various manifestations.^22^ These analyses provide a more nuanced understanding of the genetic architecture of the disorder by exploring whether certain genetic pathways implicated in schizophrenia also relate to potential retinal changes. This could help elucidating the biological mechanisms underlying both the psychiatric and retinal manifestations of schizophrenia. For instance, if certain pathways are associated with both higher genetic risk for schizophrenia and specific retinal changes, this could indicate a shared biological basis that contributes to both the psychiatric symptoms and the observed ophthalmic abnormalities. In summary, we hypothesized that the thickness of the macula in individuals with a greater polygenic risk for schizophrenia would decrease while controlling for confounding factors, and that this association would be reflected in pathways relevant for schizophrenia.

**Figure 1.**
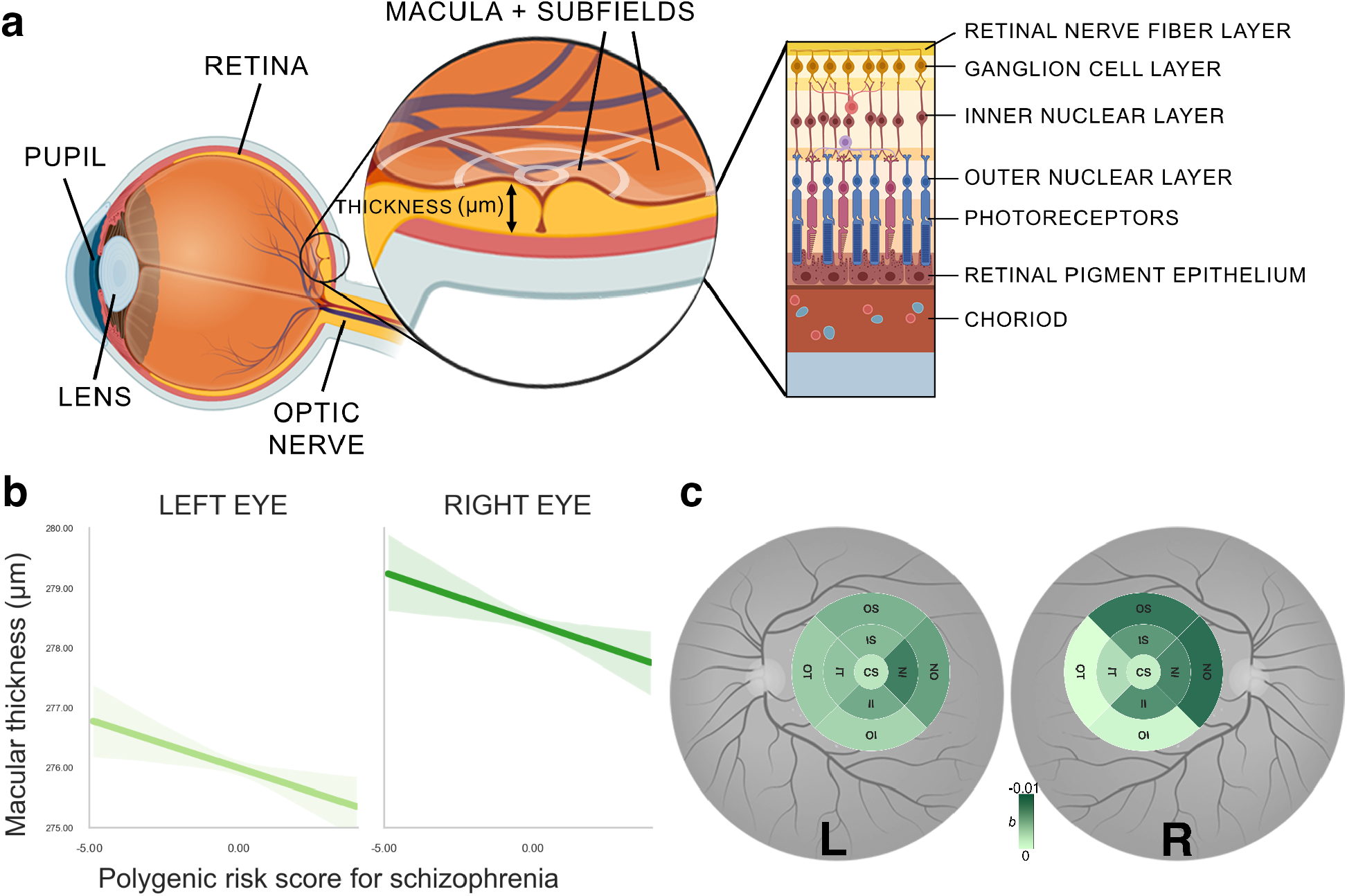
a. Illustration of the anatomy of the eye and retina, displayed as the cross section of the eye, and macular section of the retina. Optical coherence tomography measurements divide the macula in 9 subfields as indicated by white crosshairs. Macular thickness is measured in micrometers (μm). Structure of different retinal layers is displayed on the right. b. Association between polygenic risk score for schizophrenia and overall macular thickness in each eye respectively. Solid lines represent the regression estimates, while complementary shaded areas correspond to 95 percent confidence intervals. c. Individual macular subfields associated with polygenic risk scores for schizophrenia for both left eye (left) and right eye (right). Color coding corresponds to coefficients (b) from robust linear regression model. Higher b values correspond to greater thinning with increasing polygenic risk scores for schizophrenia. The sizes of subfields are slightly inflated for visualization purposes only. CS: Center subfield, IS: Inner Superior, OS: Outer Superior, IN: Inner Nasal, ON: Outer Nasal, II: Inner Inferior, OI: Outer Inferior, IT: Inner Temporal, OT: Outer Temporal.

## Methods

### Base data: schizophrenia genome-wide association study (GWAS) 2022

The summary statistics base dataset was derived from the latest available genome-wide association study for schizophrenia.^11^ We used a file generated with the exclusion of samples from the UK Biobank to assure that the base and discovery files are independent. Following quality control recommendations,^12^ we assured the same genome build with the discovery data (GRCh37/hg19), retained SNPs with minor allele frequency > 1% and INFO score > 0.8, checked for duplicate SNPs and removed ambiguous single-nucleotide polymorphisms (SNPs). The final base file included 5,899,135 SNPs.

### Discovery data: UK Biobank genetic dataset

This study used data from the UK Biobank (application 102266). A full description of genotyping and imputation procedures of the UK Biobank data (https://www.ukbiobank.ac.uk/) is provided in the release documentation elsewhere.^23^ Briefly, 487,409 blood samples were assayed using two customized tagSNP arrays (the Applied Biosystems UK BiLEVE Axiom Array and the Applied Biosystems UK Biobank Axiom Array [Affymetrix, Santa Clara, CA, USA]) with 95% shared markers, imputed to the UK10K and 1,000 Genome Project Phase 3 reference panels, with SHAPEIT3^24^ used for phasing, and IMPUTE2^25^ used for imputation. Further data handling and QC steps were carried out according to a published processing pipeline.^26^ To address population stratification, we retrieved ten genetic principal components from the UK Biobank. Specifically, after SNP extraction and alignment, conversion from bgen to PLINK format, removal of ambiguous SNPs (A/T, C/G; effects with allele frequencies between 0.4 and 0.6), data underwent a SNP-level QC (MAF < 0.005 and INFO score < 0.4) and sample-level QC (retaining individuals with missing rate in autosomes=< 0.02, which were not outliers for genotype missingness or heterozygosity, not being genetically related up to third-degree relatives, not being sex-discordant and of White British or Irish ethnicity according to genetic grouping). We also excluded individuals with ICD-10 diagnosis (F20 to F29), those who were medicated with antipsychotics and had missing data in the variables of interest or covariates. In total, n = 48208 individuals with matched imaging-genetic data were included in the final analysis (see Figure 2 for details).

**Figure 2.**
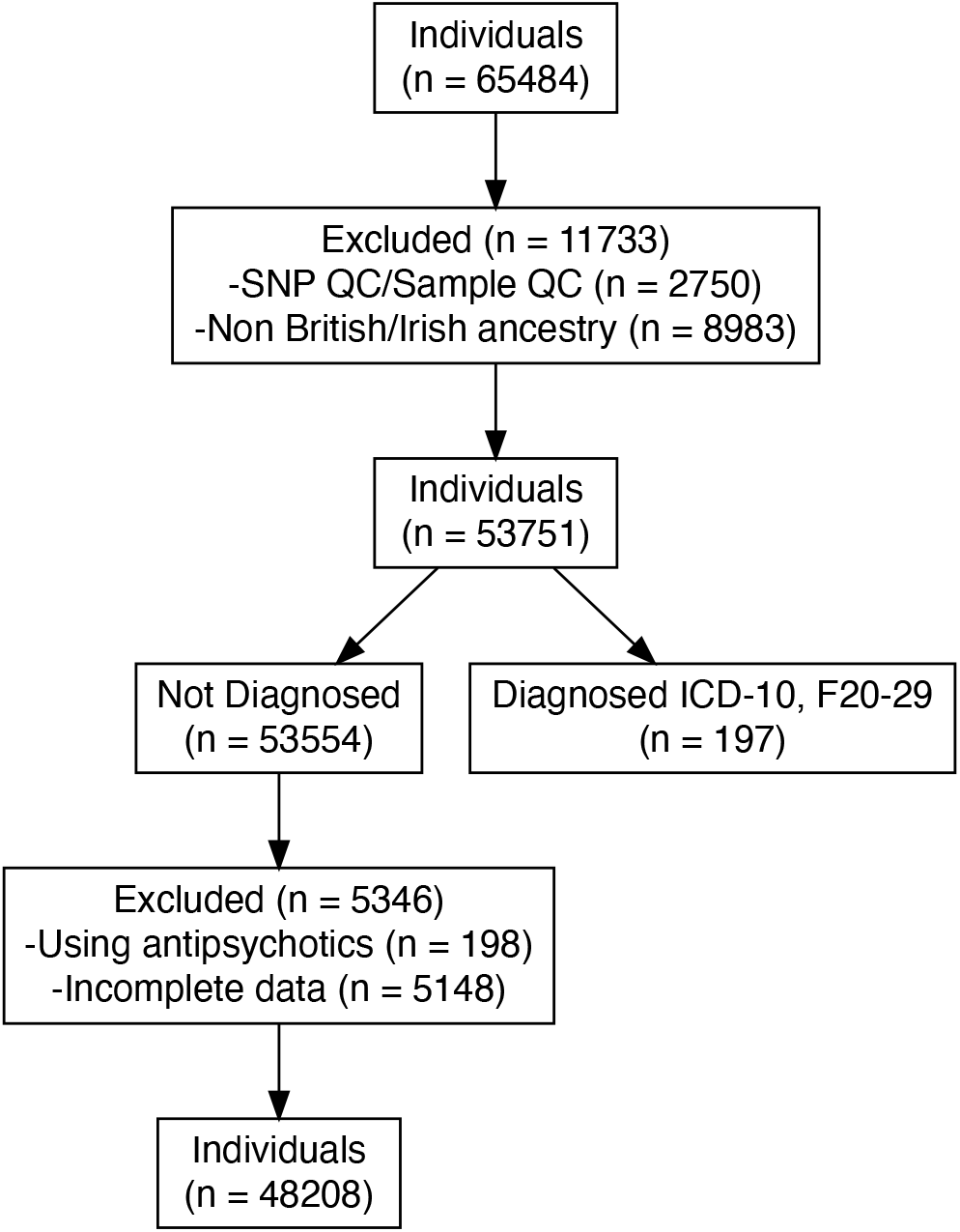
Diagram of inclusion and exclusion of participants in population. QC = Quality control. ICD = International Classification of Diseases. ICD-10, F20-29 categorization includes individuals with diagnosed schizophrenia, schizotypal and delusional disorders.

### Polygenic risk scores calculations

In the main analysis, polygenic risk scores for schizophrenia were computed for each individual as a sum of risk alleles weighted by their estimated effect sizes^27^ using the –score function in PLINK 2.0.^28^ In addition, we generated five polygenic risk scores for schizophrenia for each individual, employing SNPs selected based on their significance in association with the phenotype in the discovery GWAS at nominal p-value thresholds of 0.01 or less, 0.05, 0.1, 0.5, and 1.00.

### Derivation of pathway-based polygenic risk scores and signaling-pathway-specific analyses

We prioritize pathways known to be associated with the disease or phenotype of interest based on previous studies.^18–21^ Pathway polygenic risk scores were calculated by including only those SNPs that were relevant to the specific pathway under investigation and are also associated with schizophrenia. Pathway polygenic risk scores for schizophrenia were computed using PRSet^12^ in PRSice-2^27^ for nine candidate gene sets selected from the Molecular Signatures Database version v2023.2 (https://www.gsea-msigdb.org/): acute inflammatory response (systematic name: M6557), TGF-beta signaling (M18933), chronic inflammatory response (M15140), positive regulation of dopamine receptor signaling pathway (M24111), Wnt signaling pathway involved in midbrain dopaminergic neuron differentiation (M25305), Wnt/beta-catenin pathways (M17761), neuroinflammatory response (M24927), abnormal retinal vascular morphology (M43559) and premature coronary artery atherosclerosis (M36658). For more details, see *Gene pathways from the Molecular Signatures Database (MSigDB) version 7.4 utilized in pathway-based analysis* in the Supplementary Information. A PRSet p-value threshold was set at 1 due to the limited number of SNPs in gene-set polygenic risk scores, potentially not reflecting the entirety of gene sets accurately. Both the self-contained p-value and the competitive p-value were obtained. Self-contained methods tested each gene set independently to determine if the genes within the set were associated with the phenotype of interest. This approach does not compare the gene set to the rest of the genome. The level of association reflected by self-contained methods is the degree to which the genes within the pathway are collectively associated with the phenotype, without considering genes outside the pathway.^12^ Competitive methods, on the other hand, compared the level of association between genes within a pathway and the rest of the genes in the genome. This approach tested whether the genes in the pathway are more associated with the phenotype than what would be expected by chance, given the level of association observed in genes outside the pathway. Thus, this competitive method reflected signal enrichment by determining if the pathway stood out against the genomic background.^12^ Both self-contained and competitive p-values were calculated within PRSet through 10,000 permutations to generate null distribution curves for p-values. The regression models consisted of either initial PC1 or PC2 and all covariates. Human GRCh37 genome version was used as the background file.

### Optical coherence tomography protocol and analysis

Optical coherence tomography images were acquired using a spectral domain optical coherence tomography device, with a raster scan protocol of 6×6mm area centered on the fovea, consisting of 128 B-scans each with 512 A-scans, completed in 3.7 seconds. Automated analysis of retinal thickness was performed using custom software developed by Topcon Advanced Biomedical Imaging Laboratory, which used dual-scale gradient information for rapid segmentation of nine intraretinal boundaries, processing the images in approximately 120 seconds each. A comprehensive account of the standardized protocol employed for optical coherence tomography acquisition and the subsequent automated analysis of retinal thickness has previously been described.^29^ In the current study, we focused on the macula as it contains mutiple layers that based on prior investigations show thinning in individuals diagnosed with schizophrenia.^9,30,31^

### Assessments of optical coherence tomography data distribution and collinearity

The normality of the distribution for each retinal phenotype was assessed by visual inspection. However, the tailedness of the distributions for each retinal phenotype data appeared to be skewed by outliers (see diagonal in Figure S3). The presence of heteroscedasticity in linear regression models was evaluated using the Breusch-Pagan test. To assess collinearity, we calculated the variance inflation factor for each macular subfield, with a value greater than five indicating moderate and greater than ten indicating excessive or serious multicollinearity (Table S3). Pearson correlation coefficients between retinal phenotypes were computed (Figure S3) and visualized as a heatmap (Figure S1).

### Robust linear regression analysis for overall macular thickness and polygenic risk scores for schizophrenia

To account for potential outliers and heteroscedasticity observed in the retinal phenotype data, robust regression analysis was employed in order to examine the association between the polygenic risk scores for schizophrenia and macular phenotypes.

Robust linear regression employs M-estimation for robust linear modeling. M-estimators are a broad class of estimators in statistics that generalize maximum likelihood estimators, which are sensitive to outliers and violations of normality assumptions. The estimator used a Huber weight-function to downweight the influence of outliers and heavy-tailed distributions on the estimation of the model parameters.^32^ We used the MASS::rlm package in R to conduct such a robust analysis consisting of overall macular thickness as independent and polygenic risk scores for schizophrenia as dependent variables.

In this study when reporting *b*, we always refer to the standardized regression coefficient, which represents the estimated change in the dependent variable for a one-unit change in the independent variable.

### Confounding factors

We included a comprehensive set of covariates in a multiple linear regression analysis for studying the association between polygenic risk scores for schizophrenia and macular changes. These were age and quadratic age terms, sex, the type of genotyping array used, Body Mass Index (BMI), smoking status, optical coherence tomography image quality (OCT quality), Macula centeredness, eye disorders, Townsend Deprivation Index, genotype array, and the first ten genetic principal components. The following rationale was applied for the inclusion of covariates: Age is a fundamental factor in the development of diseases, including macular changes and schizophrenia.^33^ Various retinal structures are also known ti degenerate with age.^34^ The inclusion of both linear and quadratic terms for age allows the model to capture not just a linear increase or decrease in risk or severity with age, but also any acceleration or deceleration in this trend. Furthermore, biological sex can influence the risk of developing various diseases, their progression, and response to treatment,^35^ and there is some evidence for sex differences on some retinal structural parameters.^36^

BMI is a well-known risk factor for a wide range of health conditions, including eye diseases.^33^ It can influence the development and progression of macular changes. Similarly, smoking has been linked to an increased risk of various diseases, including those affecting the eyes.^35^

Different genotyping arrays might have varying levels of accuracy or might target different sets of genetic variations. Likewise, the quality of optical coherence tomography images and the positioning of the macula in the images can influence the ability to detect macular changes.

The Townsend Deprivation Index in UK Biobank reflects socioeconomic status, which can influence health outcomes, including those related to ophthalmic health.^35^ We also controlled for eye disorders and diseases known to affect the eye, including diabetetic retinopathy, glaucoma, macular degeneration, injury or trauma resulting in loss of vision.

Finally, the first ten genetic principal components account for population stratification, which can confound genetic associations. Including them in our models helps to ensure that any associations found are not due to underlying population genetic differences.^33^

### Principal component analysis

To account for anticipated collinearity among macular subfields measures, we conducted a principal component analysis (PCA) to obtain orthogonal principal axes reflecting variations in macular subfields. Each principal axis represents a linear combination of the original macular subfields. Prior to conducting principal component analysis, we adjusted each phenotype for the confounding variables. The resultant model residuals were then standardized and used for principal component analysis. We selected the first two principal components (columns) that collectively accounted for at least 70% of the variance in the macular data.^37^ These principal components were then used in principal component regression. To aid in the interpretation of the principal components, we displayed the direction and strength of each retinal phenotype’s contribution to a specific principal axis included in the regression model. This visualization helped to understand the relationships between the retinal phenotypes and the principal components (Figure S2).

### Robust principal component regression

Similarly, we also computed robust regression analyses consisting of either the first or second principal components as independent and and polygenic risk scores for schizophrenia as dependent variables. We not only extended this analysis to polygenic risk scores based on the discovery GWAS at different nominal p-value thresholds, but also to pathway-specific polygenic risk scores. Regression coefficients were computed, and each regression model underwent a robust F-test to determine statistical significance using sfsmisc::f.robftest package. To account for multiple comparisons, adjusted p-values (pFWER) were calculated to correct for family-wise error rate (FWER).

### Statistical thresholding

Statistical significance for individual analyses was defined as p < 0.05. The pFWER significance threshold using Holm-Bonferroni method was set at pFWER < 0.1.

### C-reactive protein (CRP)

The UK Biobank has meticulously recorded an array of biomarkers, with comprehensive details about their storage and analysis available at the Biobank showcase. The measurement of serum CRP levels was carried out using a high-sensitivity immunoturbidimetric method on a Beckman Coulter AU5800 analyzer. In our analysis, we applied a logarithmic transformation to the CRP levels to address their notably skewed distribution, as illustrated in Figure S4.

### Partial effect analyses

In order to visualize individual phenotypes, we computed the correlation coefficients between the polygenic risk scores for schizophrenia and overall macular thickness for each eye respectively while regressing out all confounding factors.

### Mediation analysis

We further sought to elucidate the mechanisms underlying the association between a pathway-enriched polygenic risk score for schizophrenia, specifically the neuroinflammatory pathway, and principal component 1. Moreover, we investigated the potential mediating role of CRP in this relationship. The rationale for examining CRP as a mediator is grounded in the evidence linking neuroinflammatory processes to the pathophysiology of schizophrenia.^38,39^ Elevated levels of CRP have been associated with increased risk and severity of schizophrenia, suggesting that inflammation may be a biological pathway through which genetic risk factors exert their effects on retinal phenotypes. To assess the mediation effect, we conducted a robust mediation analysis using two models. The mediator model was a robust linear regression that predicted CRP levels from neuroinflammatory-enriched PRS while controlling for before mentioned covariates. This model allowed us to estimate the effect of neuroinflammatory PRS on CRP (Path A). The outcome model was another robust linear regression that predicted PC1 from both neuroinflammatory PRS and CRP, controlling for the same co-variates. This model provided estimates for the direct effect of neuroinflammatory PRS on PC1 (Path C) and the effect of CRP on PC1 while accounting for neuroinflammatory PRS (Path B).

The indirect effect (Path C’ = Path A * Path B), representing the mediation effect of CRP, was calculated as the product of the coefficients from paths a and b. To evaluate the significance of the effect, we employed a bootstrap method with 1000 resamples, generating empirical confidence intervals for the mediation effect. This non-parametric approach allowed us to infer the robustness of the mediation effect without relying on the assumptions of normality. The p-value associated with this effect was computed through confidence interval inversion.

### Data and code availability

All data utilized in this study is publicly available at the UK Biobank (http://www.ukbiobank.ac.uk/).

All data analyses were performed between June 2023 and Feburary 2024 using Python (version 3.9.12) and R (version 4.0.5). The manuscript was produced with the R packages rmarkdown (version 2.21); MASS (version 7.3.58.4); SjPlot (version 2.8.14); sfsmisc (version 1.1.16); represearch (version 0.0.0.9000; https://github.com/phoman/represearch/); knitr (version 1.43); and papaja (version 0.1.1). All code will be made freely available after publication to ensure reproducibility at https://github.com/homanlab/retinflam/.

## Results

### Overall reporting details

Out of the 65484 individuals recruited with available genetic and retinal data, 48208 individuals (73.60 %) were included for further analysis. Among these participants, (53.50 %) were females, and (46.50 %) were males (Table 1). We excluded 5543 individuals due to incomplete optical coherence tomography imaging for all phenotypes, the use of antipsychotics medications and and a diagnosis ICD-10 (F20-F29; see Figure 2 for all details).

**Table 1.** Sample characteristics.

| Characteristic | N | Mean (SD) |
| --- | --- | --- |
| Female | 25781 | - |
| Male | 22427 | - |
| Eye disorder | 2099 | - |
| Current smoker | 4502 | - |
| Previous smoker | 17591 | - |
| Non smoker | 26115 | - |
| Age | 48208 | 57.83 (7.96) |
| BMI | 48208 | 27.32 (4.71) |
| Townsend Index | 48208 | -1.33 (2.81) |
| Overall macular thickness (left) | 48208 | 274.52 (25.92) |
| Overall macular thickness (right) | 48208 | 277.23 (23.86) |
| Macular thickness at the inner inferior subfield (left) | 48208 | 306.29 (32.58) |
| Macular thickness at the outer inferior subfield (left) | 48208 | 261.73 (30.08) |
| Macular thickness at the inner nasal subfield (left) | 48208 | 314.52 (32.09) |
| Macular thickness at the outer nasal subfield (left) | 48208 | 286.75 (27.68) |
| Macular thickness at the inner superior subfield (left) | 48208 | 307.65 (35.44) |
| Macular thickness at the outer superior subfield (left) | 48208 | 264.25 (29.97) |
| Macular thickness at the inner temporal subfield (left) | 48208 | 297.41 (33.39) |
| Macular thickness at the outer temporal subfield (left) | 48208 | 248.72 (28.08) |
| Macular thickness at the central subfield (left) | 48208 | 267.33 (35.23) |
| Macular thickness at the inner inferior subfield (right) | 48208 | 308.0 (32.77) |
| Macular thickness at the outer inferior subfield (right) | 48208 | 262.48 (29.35) |
| Macular thickness at the inner nasal subfield (right) | 48208 | 314.16 (32.79) |
| Macular thickness at the outer nasal subfield (right) | 48208 | 282.49 (27.94) |
| Macular thickness at the inner superior subfield (right) | 48208 | 309.83 (34.24) |
| Macular thickness at the outer superior subfield (right) | 48208 | 268.97 (29.17) |
| Macular thickness at the inner temporal subfield (right) | 48208 | 300.44 (32.2) |
| Macular thickness at the outer temporal subfield (right) | 48208 | 258.17 (27.64) |
| Macular thickness at the central subfield (right) | 48208 | 269.56 (36.19) |

The overall macular thickness appeared to be significantly larger in the right eye (M = 277.23) compared to the left eye (M = 274.77; t = 24.45, p < .001).

In the analysis of the relationship between polygenic risk scores for schizophrenia and retinal phenotypes, the linear regression models displayed heteroscedasticity, as evidenced by all Breusch-Pagan tests yielding significant results (p<0.001, Table **??**). This indicates that the variance of the residuals was not constant across the range of values, suggesting that the precision of the regression estimates varies depending on the level of the polygenic risk scores.

### Polygenic risk for schizophrenia can predict thinning of the macula

The primary dependent variable of the study was the thickness of the macula and that of its nine subfields (Figure 1a).

We conducted robust regression analyses to examine the relationships between polygenic risk scores for schizophrenia and overall macular thickness for each eye respectively. For the left eye, we observed a negative relationship between macular thickness and polygenic risk scores for schizophrenia (b = −0.16, SE = 0.06, p = 0.009, pFWER = 0.02; Figure 1b). This means that for each one standard deviation increase in polygenic risk for schizophrenia, the thickness of the left macula decreased by 0.16 micrometers. We also found a similar negative association for the right eye (b = −0.17, SE = 0.06, p = 0.009, pFWER = 0.02; Figure 1b).

Additionally, we computed robust regression analyses for each macular subfield separately (Figure 1c). More specifically, superior and nasal subfields showed the greatest thinning in both eyes (left OS: b = −0.01, p = 0.012, pFWER = 0.15 right OS: b = −0.01, p < .001, pFWER = 0.01, left ON: b = −0.01, p = 0.020, pFWER = 0.18, left IN: b = −0.01, p = 0.003, pFWER = 0.04, right ON: b = −0.01, p < .001, pFWER = 0.00). In summary, these results suggest that with increasing polygenic risk for schizophrenia, the overall macula becomes thinner, and within each eye, macular subfields seem to be thinning non-uniformly. However, testing the latter proved to be difficult due to the high collinearity between thickness measures of different macular subfields (Figure S1).

Therefore, we reduced by means of principal component analysis the dimensionality of data by transforming original retinal phenotypes into a smaller number of principal components that explain most of the variance in the data. Compared to our overall macular thickness measures, this reduction in dimensionality and regressing out of the confounding variables improves the statistical power of genetic association testing by reducing the multiple testing burden and highlighting the most relevant variation for disease risk.^40^ The first two principal components explained 72 % of the data variance (Figure S2a). While principal component 1 captured a pattern of variation characterized by a general increase of thickness across macular subfields, principal component 2 captured a pattern of variation characterized by an increase of macular subfields of the left eye and a decrease of the right eye (Figure S2b). In a robust linear regression analysis, we examined the correlation coefficients for each principal component contributing to this multivariate model. We observed a negative association between the polygenic risk scores for schizophrenia and principal component 1 (b = −0.03, SE = 0.01, p = 0.007, pFWER = 0.01). This means that for each one standard deviation increase in polygenic risk for schizophrenia, the value of the first principal component tends to decrease by 0.03 standard deviations, indicating that the first principal component captures a dimension of the data that is negatively correlated with polygenic risk scores for schizophrenia. As expected, this association was consistent with individual regression results showing a negative relationship between polygenic risk scores for schizophrenia and overall macular thickness. The association between polygenic risk scores for schizophrenia and principal components 2 was not significant (b = 0, SE = 0.00, p = 0.306, pFWER = 0.31). This suggests that the specific pattern of macular thickness differences between the left and right eye as captured by the second component may not be a relevant biomarker for predicting the genetic risk for schizophrenia.

### Neuroinflammatory pathway-specific polygenic risk scores are associated with macular thinning

Pathway-specific polygenic risk scores provided us with a more nuanced understanding of the genetic contribution to genetic risk manifestations. Intriguingly, we also observed that polygenic risk scores specific to neuroinflammation (b = −0.03, self-contained p = 0.041, competitive p = 0.051) and Wnt signalling pathways involved in midbrain dopaminergic neuron differentiation (b = −0.02, self-contained p = 0.063, competitive p = 0.079) in relation to schizophrenia revealed a significant association and a trend to significance with the first principal component, respectively (Figure 3). We found no statistically significant associations with other gene pathways that potentially could be related to schizophrenia (for more details, see Table S2). Thus, genetic variation in developmental and inflammatory pathways may confer risk for schizophrenia through effects on macular thickness.

**Figure 3.**
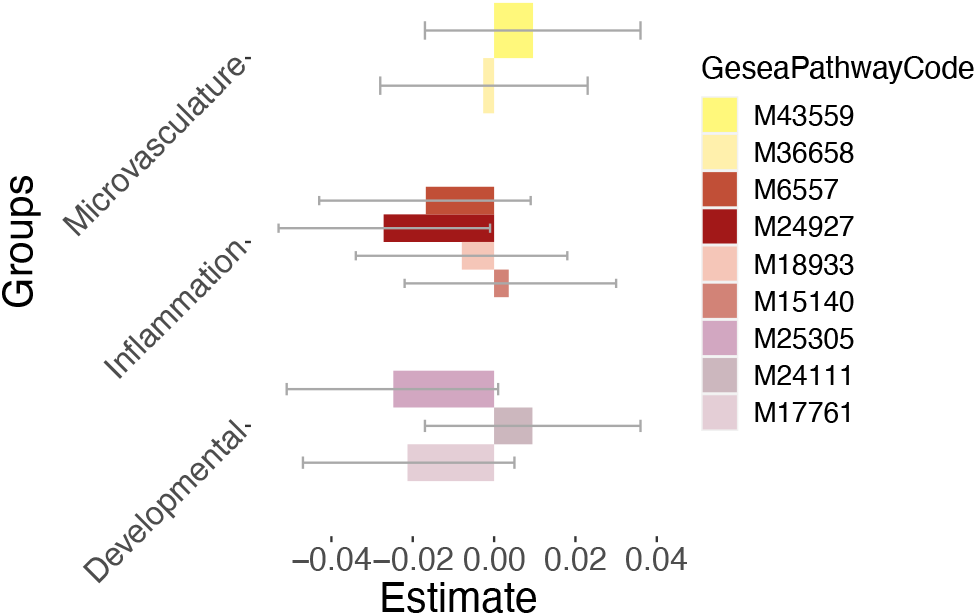
Association between polygenic risk for schizophrenia enriched for multiple gene pathways and first retinal principal component at p-value threshold 1. Confidence intervals (95 percent) are visualized by grey error bars. Microvasculature pathways: 1. HP abnormal retinal vascular morphology (M43559) 2. HP premature coronary artery atherosclerosis (M36658). Inflammatory pathways: 1. GOBP acute inflammatory response (M6557) 2. GOBP neuroinflammatory response (M24927) 3. Biocarta TGFB pathway (M18933) 4. GOBP chronic inflammatory response (M15140). Signaling pathways influencing neuronal development: 1. GOBP Wnt signaling pathway involved in midbrain dopaminergic neuron differentiation (M25305) 2. GOBP positive regulation of dopamine receptor signaling pathway (M24111) 3. ST Wnt beta catenin pathway (M17761).

### CRP levels partially mediate the association between neuroinflammatory pathway-specific PRS and retinal thickness

The UK Biobank also provided us with CRP measurements, a protein that plays a pivotal role in managing inflammation. Therefore, we explored the mediating role of CRP in the relationship between neuroinflammatory-enriched polygenic risk scores for schizophrenia and the principal component 1 as an outcome variable (Figure 4). The mediator model revealed a significant effect of neuroinflammatory-specific polygenic risk scores for schizophrenia on CRP (Path A coefficient = 0.01), controlling for confounding factors. This positive correlation between neuroinflammation gene enriched polygenic risk scores for schizophrenia and CRP levels suggests that individuals with a higher genetic risk for schizophrenia through neuroinflammatory-related genes also have higher systemic inflammation, as indicated by CRP levels. Furthermore, the outcome model showed a significant effect of CRP on PC1 (Path B coefficient = −0.07), after accounting for the same covariates and neuroinflammatory polygenic risk for schizophrenia. The estimated indirect effect of neuroinflammatory-specific polygenic risk for schizophrenia on PC1 through the mediator CRP was −0.001, with a 95% bootstrap confidence interval of [−0.002, −0.0003], suggesting a statistically significant partial mediation effect (p = 0.001). This means that part of the negative impact of neuroinflammatory-specific polygenic risk scores for schizophrenia on PC1 is mediated by increased CRP levels.

**Figure 4.**
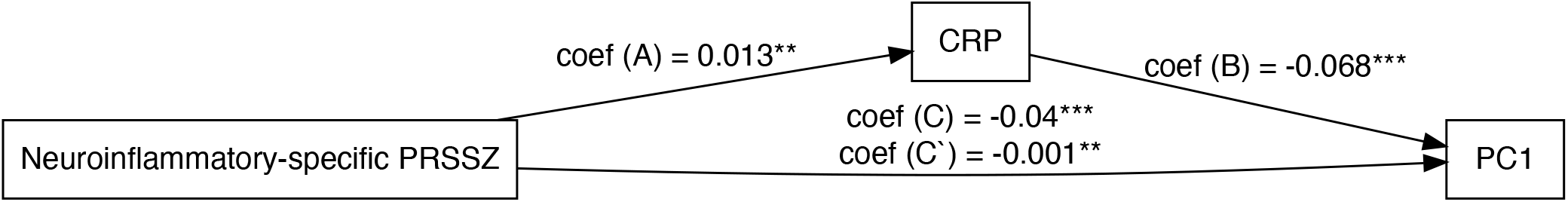
Mediation analysis: C-reactive protein related to associability acting as a partial mediator in the link between polygenic risk scores specific to neuroinflammation for schizophrenia and macular phenotypes represented by PC1. Coef = beta coefficient, PRSSZ = polygenic risk for schizophrenia, CRP = C-reactive protein. ** = p < 0.01, *** = p < 0.001

## Discussion

In this large observational study, we discovered that an increased genetic predisposition to schizophrenia is associated with retinal thinning. More specifically, we observed structural abnormalities of the macula in individuals with higher polygenic risk scores for schizophrenia. Our findings also indicate potential involvement of gene variants related to neuroinflammatory pathways in the expression of retinal phenotypes.

To the best of our knowledge, this is the first study to explore the association between genetic risk for schizophrenia and retinal variations within a large population-based sample using the latest GWAS.^11^ Moreover, our results extend prior findings of retinal thinning in patients with schizophrenia compared to healthy individuals,^4,5,8,9,31^ demonstrating an association between genetic risk for schizophrenia and these retinal phenotypes.

Our population data highlights an asymmetry of macular thickness between the left and right eyes, which has been reported before.^41^ However, this asymmetry did not appear to play a major role in which eye is more affected by a higher genetic risk for schizophrenia. Within each eye, the limited number of principal components did not allow us to discern differences in thinning between different macular subfields. Therefore, whether different diseases target specific anatomical structures of the retina, e.g. only ganglion cells, thus preferentially affecting subfields with a higher density of ganglion cells stronger than others is a question in need of further research. Understanding these differences has implications for using retinal imaging data as an aid to diagnosis and patient monitoring during treatment. Although we provided evidence of a genetic link between polygenic risk scores for schizophrenia and retinal thinning, the exact mechanisms underlying these retinal alterations remain unclear. The retina is susceptible to systemic neurodegenerative processes that also affect the brain. We therefore explored different genetic pathways that could potentially affect the retina in individuals with increasing polygenic risk for schizophrenia. Our findings suggest an association between not only neuroinflammatory pathways and retinal thickness but also Wnt signaling pathways for midbrain dopaminergic neuron differentiation. Inflammatory processes can disrupt the normal functioning of astrocytes, potentially leading to neurotransmitter dysregulation and blood–brain barrier permeability.^42,43^ Blood-brain barrier disruption, caused by pro-inflammatory cytokines and chemokines, can exacerbate neuroinflammation by allowing immune cells and potentially harmful substances to enter the brain.^42,44^ This can lead to the release of acute-phase proteins, oxidative stress, excitotoxicity, and other processes that cause neuronal damage, affecting synaptic functioning, which is critical for normal cognitive processes and has been found to be disrupted in schizophrenia.^42,44^ Acute-phase proteins like CRP can also contribute to the progressive apoptosis of photoreceptors,^45,46^ which might explain that our finding of a partial mediation effect of CRP, suggesting its mediating role next to other potential candidates, on the association between neuroinflammatory enriched polygenic risk scores for schizophrenia and macular thickness. This suggests that neuroinflammatory processes, as indexed by CRP levels, could be a key biological pathway through which genetic risk factors for schizophrenia influence macular thickness.

While a previous study found no association between the levels of inflammatory markers (CRP among others) and retinal layer changes in either the schizophrenia or control group,^47^ that study was considerably smaller than the present study. It is possible that such an effect can only be detected with substantially more statistical power, coming from a large sample size as in our study.

Furthermore, the interplay between neuroinflammatory and Wnt signaling pathways is critical in regulating the immune response within the CNS,^48,49^ which might explain our second pathways-specific finding. The Wnt signaling pathway is crucial for normal brain development, including the differentiation of dopaminergic neurons in the midbrain, which are implicated in the pathophysiology of schizophrenia.^21^ If the Wnt signaling pathway is altered in individuals with a higher genetic risk for schizophrenia, this could lead to developmental abnormalities in the retina, a structure that shares embryological origins with the brain. Such abnormalities could manifest as changes in retinal thickness. This might explain its association with macular thinning in individuals with higher polygenic risk scores for schizophrenia in our study. However, this result warrants a cautious interpretation since the association only approached significance. Nonetheless, our findings on macular thinning could potentially reflect Wnt dysregulation which can lead to deleterious effects in neuronal development, contributing to the pathogenesis of neurodevelopmental diseases like schizophrenia.^19,50^ Other Wnt signaling pathways also play a significant role in the protection of damaged retinal neurons.^51^ They have been shown to have a significant impact on retinal vessel formation and maturation, as well as on the establishment of synaptic structures and neuronal function in the central nervous system.^51^ The activation of certain Wnt pathways has been shown to reverse the pathological phenotype caused by Wnt inhibition in the context of glaucoma.^52^ However, our other Wnt signaling pathways did not show any significant association with macular thickness changes. This could have occured because our selection of these pathways was very limited and/or the gene sets of these pathways did not fully capture the polygenic nature of the effects.

In summary, the genetic risk factors for schizophrenia, as represented by the polygenic risk scores, may influence neuroinflammatory and Wnt signaling pathways involved in the disease. These pathways could, in turn, affect the neural and vascular integrity of the retina, leading to observable retinal changes. However, while we observed a relationship between polygenic risk scores and neural layer thinning at the macula, we did not observe any associations between retinal microvasculature gene-based polygenic risk scores for schizophrenia and retinal phenotypes. This could be due to the fact that genetic architecture of retinal microvasculature traits and schizophrenia risk is complex and again, the selected pathways were not able to capture this relationship.

Alternatively, retinal thinning in individuals with high polygenic risk for schizophrenia may also be influenced by other factors e.g. systemic comorbidities like hypertension, metabolic dysfunction, certain health behaviors, and life-course exposures, which were not evaluated in this study. Therefore, conclusive evidence for underlying processes in schizophrenia are subject to further investigations.

Several limitations merit comment. The relatively high mean age in our study may not fully represent younger individuals with higher polygenic risk for schizophrenia. Moreover, the UK Biobank sample may not be fully representative of the UK population, and our focus on White British or Irish participants limits generalizability to other ethnicities. Additionally, the polygenic risk score’s explanatory power for schizophrenia is modest, which may have limited the detection of small effects. Our study also had several strengths, including the use of a large, population-based sample from the UK Biobank, which allowed us to avoid confounding factors related to psychotic and retinal illnesses or antipsychotic use. We also employed robust statistical analyses and complementary techniques to enhance the reliability of our results. Additionally, our extended analyses produced consistent findings across multiple inclusion thresholds for the polygenic risk score.

In conclusion, our study identified significant associations between polygenic risk scores for schizophrenia, pathway-specific scores, and retinal phenotypes, offering valuable insights into the genetic contributions to retinal changes in individuals without any diagnosis of a psychotic disorder or schizotypal personality disorder. As genome-wide association studies expand, and as the polygenic risk score for schizophrenia is updated future research may reveal more subtle associations between polygenic risk scores for schizophrenia and retinal phenotypes.

## Acknowledgements

Figures 1a and 1c include graphics created with BioRender.com.

## Author contributions

FR: conceptualization, methodology, statistical analysis, visualization, writing, review and editing. LK: methodology, genetic analysis, writing, review and editing. FG: conceptualization, methodology and statistical analysis. NK: conceptualization and supervision. WO: review and editing. MK: review and editing. FC: review and editing. EG: methodology, review and editing. SS: supervision, review & editing. BB: review & editing. DB: review & editing. KS: review & editing. JR: review & editing. TL: supervision, methodology, review & editing. PH: supervision, conceptualization, writing, review and editing.

## Supplementary Information

### Supplementary Results

**Gene pathways from the Molecular Signatures Database (MSigDB) version 7**.**4 utilized in pathway-based analysis. GOBP_NEUROINFLAMMATORY_RESPONSE** > Neuroinflammatory response is the nervous system’s reaction to various forms of damage, including infection, traumatic brain injury, toxic metabolites, or autoimmunity. [ADCY1 ADCY8 ADORA2A AGER AIF1 APP ATM AZU1 BPGM C1QA C5AR1 CCL3 CD200 CD200R1 CD200R1L CLU CNTF CST7 CTSC CX3CL1 CX3CR1 DAGLA GNAT2 GRN IFNG IFNGR1 IFNGR2 IGF1 IL13 IL18 IL1B IL33 IL4 IL6 ITGAM ITGB1 ITGB2 JAK2 KCNJ8 LARGE1 LDLR LRP1 LRRK2 MAPT MIR128-1 MIR142 MIR181B1 MIR181C MIR195 MIR206 MIR26A1 MMP3 MMP8 MMP9 NAGLU NR1D1 NR3C1 NUPR1 PLCG2 POMGNT1 POMT2 PSEN1 PTGS2 PTPRC SMO SNCA SPHK1 STAP1 SYT11 TAFA3 TLR2 TLR3 TLR6 TLR9 TNF TNFRSF1B TREM2 TRPV1 TTBK1 TYROBP VPS54 ZEB2]

**GOBP_CHRONIC_INFLAMMATORY_RESPONSE** > This identifier is related to the chronic inflammatory response. A chronic inflammatory response is a prolonged inflammatory reaction that can lead to tissue damage and is involved in various diseases. [ADORA2B CCL5 CXCL13 CYP19A1 FOXP3 IDO1 IL10 IL4 LTA NFKBIZ S100A8 S100A9 SCN11A THBS1 TNF TNFAIP3 UNC13D VCAM1 VNN1]

**BIOCARTA_TGFB_PATHWAY** > The TGF beta signaling pathway is a cellular mechanism that regulates various biological processes, including cell growth, differentiation, and apoptosis, and it plays a dual role in inflammation by both promoting and inhibiting inflammatory responses depending on the contex [APC CDH1 CREBBP EP300 MAP2K1 MAP3K7 MAPK3 SKIL SMAD2 SMAD3 SMAD4 SMAD7 TAB1 TGFB1 TGFB2 TGFB3 TGFBR1 TGFBR2 ZFYVE9]

**GOBP_ACUTE_INFLAMMATORY_RESPONSE** > Inflammation is characterized by a swift, transient, and relatively consistent reaction to acute harm or antigen exposure, marked by fluid, plasma protein, and granulocytic leukocyte accumulations. This acute inflammatory response, which unfolds within minutes or hours, either resolves in a few days or transitions into a chronic inflammatory state. [A2M ACVR1 ADAM8 ADORA1 ADRA2A AHSG ALOX5AP ANO6 APCS APOA2 APOL2 ASH1L ASS1 B4GALT1 BTK C2CD4A C2CD4B C3 CCR7 CD163 CD6 CEBPB CNR1 CREB3L3 CRP CTNNBIP1 DNASE1 DNASE1L3 EDNRB EIF2AK1 ELANE EPO EXT1 F12 F2 F3 F8 FCGR1A FCGR2B FCGR3A FFAR2 FFAR3 FN1 FUT7 GATA3 GSTP1 HAMP HLA-E HP HPR IGHG1 IL1A IL1B IL20RB IL22 IL31RA IL4 IL6 IL6R IL6ST INS ITIH4 KL KLKB1 LBP MBL2 MIR92A1 MRGPRX1 MYLK3 NLRP3 NLRP6 NPY NPY5R NUPR1 OGG1 OPRM1 ORM1 ORM2 OSM OSMR PARK7 PIK3CG PLA2G2D PLSCR1 PRCP PTGER3 PTGES PTGS2 REG3A REG3G RHBDD3 S100A8 SAA1 SAA2 SAA4 SELENOS SERPINA1 SERPINA3 SERPINF2 SIGIRR TACR1 TFR2 TFRC TNF TNFRSF11A TNFSF11 TNFSF4 TREM1 TRPV1 UGT1A1 VCAM1 VNN1 ZP3]

**HP_ABNORMAL_RETINAL_VASCULAR_MORPHOLOGY** > An identifier suggests a phenotype related to abnormal morphology (structure) of the blood vessels in the retina. [ABCA4 ABCC6 ACVRL1 AGBL5 AHI1 AHR AIPL1 ANTXR1 APOC2 ARHGEF18 ARL2BP ARL3 ARL6 ARV1 ARVCF ASAH1 ATF6 BAZ1B BBS1 BBS2 BBS9 BCL7B BEST1 BUD23 CA4 CAPN5 CC2D2A CCDC28B CCM2 CCND1 CDHR1 CENPF CERKL CFAP410 CFAP418 CHM CLCC1 CLCNKB CLIP2 CLRN1 CNGA1 CNGA3 CNGB1 CNGB3 CNNM4 COL18A1 COL4A1 COMT CRB1 CRX CTC1 CTNNB1 CTSA CYP1B1 CYP27A1 DARS1 DGCR2 DGCR6 DGCR8 DHDDS DHX38 DLK1 DLST DNAJC30 DNMT3A DNMT3B DUX4 DUX4L1 EFEMP1 EIF4H ELN ENG ENPP1 EPAS1 ERCC4 ESS2 ETHE1 EYS F12 FAM161A FGF12 FH FKBP6 FLVCR1 FRG1 FSCN2 FUCA1 FZD4 G6PC1 GALC GDF2 GGCX GLB1 GM2A GNAQ GNAT2 GP1BB GPIHBP1 GTF2I GTF2IRD1 GTF2IRD2 GUCA1B GUCY2D HARS1 HEXA HEXB HGSNAT HIRA HK1 HLA-A HSD11B2 IDH3A IDH3B IFT140 IFT172 IFT43 IFT88 IGFBP7 IKBKG IMPDH1 IMPG1 IMPG2 IVNS1ABP JMJD1C KIAA1549 KIF1B KIZ KLHL7 KRIT1 LAMB2 LCA5 LCK LIMK1 LOXL1 LPL LRAT LRP5 LRRC32 MAK MAX MDH2 MEG3 MERTK METTL27 MKS1 MLXIPL MT-ATP6 MT-CO1 MT-CO3 MT-CYB MT-ND1 MT-ND2 MT-ND4 MT-ND4L MT-ND5 MT-ND6 MVK MYD88 MYOC NCF1 NDP NDUFS2 NEK1 NEK2 NEU1 NF1 NMNAT1 NOD2 NR2E3 NRL NUS1 OFD1 PAX6 PCARE PCNA PDCD10 PDE6A PDE6B PDE6C PDE6G PDE6H PIK3CA POMGNT1 PRCD PROM1 PRPF3 PRPF31 PRPF4 PRPF6 PRPF8 PRPH2 PSAP RAX2 RBP3 RDH11 RDH12 RDH5 REEP6 RET RFC2 RGR RHO RLBP1 ROM1 RP1 RP1L1 RP2 RP9 RPE65 RPGR RPGRIP1 RREB1 RTL1 RTTN SAG SCAPER SDHA SDHAF2 SDHB SDHC SDHD SEC24C SELENOI SEMA4A SERPINC1 SETD2 SLC24A1 SLC25A11 SLC37A4 SLC6A6 SLC7A14 SMAD4 SMCHD1 SMPD1 SNRNP200 SPATA7 SSBP1 STN1 STX1A TBL2 TBX1 TLCD3B TMEM127 TMEM231 TMEM270 TOPORS TREX1 TSPAN12 TTC8 TUB TULP1 UFD1 UNC119 USH2A USP45 VHL VPS37D WDR19 WT1 YME1L1 ZNF408 ZNF513]

**HP_PREMATURE_CORONARY_ARTERY_ATHEROSCLEROSIS** > This term refers a gene set that is related to the premature development of atherosclerosis in the coronary arteries, which can lead to coronary artery disease (CAD). [ABCA1 ABCG5 ABCG8 ACTA2 APOA1 APOB APOE CEP19 CYP27A1 ERCC6 ERCC8 GPIHBP1 LDLR LDLRAP1 LIPC LMNA LRP6 MEF2A PCSK9]

**ST_WNT_BETA_CATENIN_PATHWAY** > This identifier refers to a gene pathway involved in the Wnt/beta-catenin signaling. The Wnt/beta-catenin pathway is crucial for various cellular processes, including development, cell proliferation, and differentiation. [AKT1 AKT2 AKT3 ANKRD6 APC AXIN1 AXIN2 CBY1 CER1 CSNK1A1 CTNNB1 CXXC4 DACT1 DKK1 DKK2 DKK3 DKK4 DVL1 FRAT1 FSTL1 GSK3A GSK3B LRP1 MVP NKD1 NKD2 PIN1 PSEN1 PTPRA RPSA SENP2 SFRP1 TSHB WIF1]

**GOBP_WNT_SIGNALING_PATHWAY_INVOLVED_IN_MIDBRAIN_DOPAMINERGIC_NEURON_DIFFERENTIATION** > This identifier contains genes that are related to the role of the Wnt signaling pathway in the differentiation of dopaminergic neurons in the midbrain. This process is important for the development of the nervous system and is particularly relevant to the study of diseases like schizophrenia and Parkinson’s disease, where dopaminergic neurons are affected. [CSNK1D CSNK1E CTNNB1 FZD1 LRP6 RYK WNT1 WNT2 WNT3 WNT3A WNT5A WNT9B]

**GOBP_POSITIVE_REGULATION_OF_DOPAMINE_RECEPTOR_SIGNALING_PATHWAY** > This identifier contains genes responsible for a biological process that positively regulates the signaling pathway of dopamine receptors. Dopamine receptors are critical for many neurological processes, and their signaling pathways are important in the context of neurological diseases, e.g. schizophrenia and Parkinsons’s disease. [CAV2 DRD3 LRRK2 PRMT5 VPS35]

### Supplementary Tables

**Table S1.** Association between polygenic risk for schizophrenia (PRSSZ) at different p-value thresholds and first two retinal principal components.

| PRS | PC | Coef | CI(low) | CI(high) | SE | F | p | pFWER |
| --- | --- | --- | --- | --- | --- | --- | --- | --- |
| PRSSZ | PC1 | -0.03 | -0.044 | -0.007 | 0.00946 | 7.26 | 0.007 | 0.0140 |
| PRSSZ 0.001 | PC2 | 0.00 | -0.006 | 0.002 | 0.00208 | 1.05 | 0.3061 | 0.3061 |
| PRSSZ 0.05 | PC1 | -0.01 | -0.029 | 0.009 | 0.0096 | 1.02 | 0.3136 | 0.3136 |
| PRSSZ 0.1 | PC2 | 0.00 | -0.007 | 0.001 | 0.00211 | 2.44 | 0.1183 | 0.2366 |
| PRSSZ 0.2 | PC1 | -0.02 | -0.042 | 0 | 0.01078 | 3.69 | 0.0547 | 0.1094 |
| PRSSZ 0.3 | PC2 | 0.00 | -0.004 | 0.005 | 0.00237 | 0.04 | 0.8383 | 0.8383 |
| PRSSZ 0.4 | PC1 | -0.02 | -0.043 | 0.001 | 0.01109 | 3.60 | 0.0578 | 0.1156 |
| PRSSZ 0.5 | PC2 | 0.00 | -0.004 | 0.006 | 0.00243 | 0.17 | 0.6821 | 0.6821 |
| PRSSZ 1 | PC1 | -0.02 | -0.042 | 0.001 | 0.0111 | 3.40 | 0.0652 | 0.1304 |
| PRSSZ | PC2 | 0.00 | -0.004 | 0.005 | 0.00244 | 0.06 | 0.8025 | 0.8025 |
| PRSSZ 0.001 | PC1 | -0.02 | -0.04 | 0.003 | 0.0112 | 2.74 | 0.0978 | 0.1956 |
| PRSSZ 0.05 | PC2 | 0.00 | -0.004 | 0.005 | 0.00246 | 0.06 | 0.807 | 0.8070 |
| PRSSZ 0.1 | PC1 | -0.02 | -0.039 | 0.005 | 0.01116 | 2.41 | 0.1202 | 0.2404 |
| PRSSZ 0.2 | PC2 | 0.00 | -0.004 | 0.006 | 0.00245 | 0.18 | 0.6751 | 0.6751 |
| PRSSZ 0.3 | PC1 | -0.02 | -0.037 | 0.007 | 0.01115 | 1.83 | 0.1757 | 0.3514 |
| PRSSZ 0.4 | PC2 | 0.00 | -0.004 | 0.006 | 0.00245 | 0.13 | 0.7233 | 0.7233 |
| PRSSZ 0.5 | PC1 | -0.02 | -0.037 | 0.007 | 0.01113 | 1.87 | 0.1714 | 0.3428 |
| PRSSZ 1 | PC2 | 0.00 | -0.004 | 0.006 | 0.00244 | 0.15 | 0.7022 | 0.7022 |

**Table S2.** Permutation test regression between polygenic risk for schizophrenia enriched for multiple gene pathways and first two retinal principal components at p-value threshold 1 using PRSet.

| Pathway | GeseaPathwayCode | Num_SNP | PC | Estimate | SE | selfcontained.p | competitive.p | Groups | selfcon.pFWER | comp.pFWER | CIhigh | CIlow |
| --- | --- | --- | --- | --- | --- | --- | --- | --- | --- | --- | --- | --- |
| ABNOVAS PRS | M43559 | 2265 | PC1 | 0.0095753 | 0.0135479 | 0.4797110 | 0.4908510 | Microvasculature | 0.959422 | 0.9817020 | 0.036 | -0.017 |
| ABNOVAS PRS | M43559 | 2265 | PC2 | 0.0040916 | 0.0074301 | 0.5818610 | 0.6275370 | Microvasculature | 0.959422 | 0.9817020 | 0.019 | -0.010 |
| ACUTEINFLAM PRS | M6557 | 817 | PC1 | -0.0167891 | 0.0133750 | 0.2093900 | 0.2312770 | Inflammation | 0.418780 | 0.4625540 | 0.009 | -0.043 |
| ACUTEINFLAM PRS | M6557 | 817 | PC2 | -0.0033672 | 0.0073354 | 0.6462180 | 0.6651330 | Inflammation | 0.646218 | 0.6651330 | 0.011 | -0.018 |
| CATENIN PRS | M17761 | 341 | PC1 | -0.0212782 | 0.0131636 | 0.1060050 | 0.1272870 | Developmental | 0.212010 | 0.2545740 | 0.005 | -0.047 |
| CATENIN PRS | M17761 | 341 | PC2 | -0.0057140 | 0.0072195 | 0.4286740 | 0.4358560 | Developmental | 0.428674 | 0.4358560 | 0.008 | -0.020 |
| CHROINFLAM PRS | M15140 | 138 | PC1 | 0.0035881 | 0.0133002 | 0.7873310 | 0.8054190 | Inflammation | 0.787331 | 0.8054190 | 0.030 | -0.022 |
| CHROINFLAM PRS | M15140 | 138 | PC2 | 0.0078411 | 0.0072942 | 0.2823910 | 0.2876710 | Inflammation | 0.564782 | 0.5753420 | 0.022 | -0.006 |
| CORART PRS | M36658 | 241 | PC1 | -0.0026740 | 0.0131740 | 0.8391530 | 0.8446160 | Microvasculature | 1.000000 | 1.0000000 | 0.023 | -0.028 |
| CORART PRS | M36658 | 241 | PC2 | 0.0036210 | 0.0072251 | 0.6162520 | 0.6251370 | Microvasculature | 1.000000 | 1.0000000 | 0.018 | -0.011 |
| DOPPOSREG PRS | M24111 | 68 | PC1 | 0.0094135 | 0.0134067 | 0.4825900 | 0.5040500 | Developmental | 0.965180 | 1.0000000 | 0.036 | -0.017 |
| DOPPOSREG PRS | M24111 | 68 | PC2 | -0.0034873 | 0.0073528 | 0.6352980 | 0.6315370 | Developmental | 0.965180 | 1.0000000 | 0.011 | -0.018 |
| NEUROINFLAM PRS | M24927 | 747 | PC1 | -0.0271559 | 0.0133030 | 0.0412220 | 0.0511949 | Inflammation | 0.082444 | 0.1023898 | -0.001 | -0.053 |
| NEUROINFLAM PRS | M24927 | 747 | PC2 | 0.0049284 | 0.0072961 | 0.4993690 | 0.5100490 | Inflammation | 0.499369 | 0.5100490 | 0.019 | -0.009 |
| TGFB PRS | M18933 | 248 | PC1 | -0.0079258 | 0.0130943 | 0.5449910 | 0.5604440 | Inflammation | 1.000000 | 1.0000000 | 0.018 | -0.034 |
| TGFB PRS | M18933 | 248 | PC2 | 0.0043773 | 0.0071814 | 0.5421730 | 0.5496450 | Inflammation | 1.000000 | 1.0000000 | 0.018 | -0.010 |
| WNT PRS | M25305 | 107 | PC1 | -0.0247761 | 0.0133031 | 0.0625475 | 0.0794921 | Developmental | 0.125095 | 0.1589842 | 0.001 | -0.051 |
| WNT PRS | M25305 | 107 | PC2 | -0.0011356 | 0.0072961 | 0.8763190 | 0.8812120 | Developmental | 0.876319 | 0.8812120 | 0.013 | -0.015 |

**Table S3.** Variance inflation factor for macular subfields in a robust linear regression model with polygenic risk scores for schizophrenia.

| Phenotype | VIF |
| --- | --- |
| Inner Inferior left | 5.60 |
| Inner Inferior right | 5.85 |
| Outer Inferior left | 3.20 |
| Outer Inferior right | 3.46 |
| Inner Nasal left | 9.33 |
| Inner Nasal right | 10.40 |
| Outer Nasal left | 6.02 |
| Outer Nasal right | 5.69 |
| Inner Superior left | 7.37 |
| Inner Superior right | 6.87 |
| Outer Superior left | 3.87 |
| Outer Superior right | 3.56 |
| Inner Temporal left | 11.42 |
| Inner Temporal right | 7.92 |
| Outer Temporal left | 5.63 |
| Outer Temporal right | 5.67 |
| Central left | 2.89 |
| Central right | 2.75 |

### Supplementary Figures

**Figure S1.**
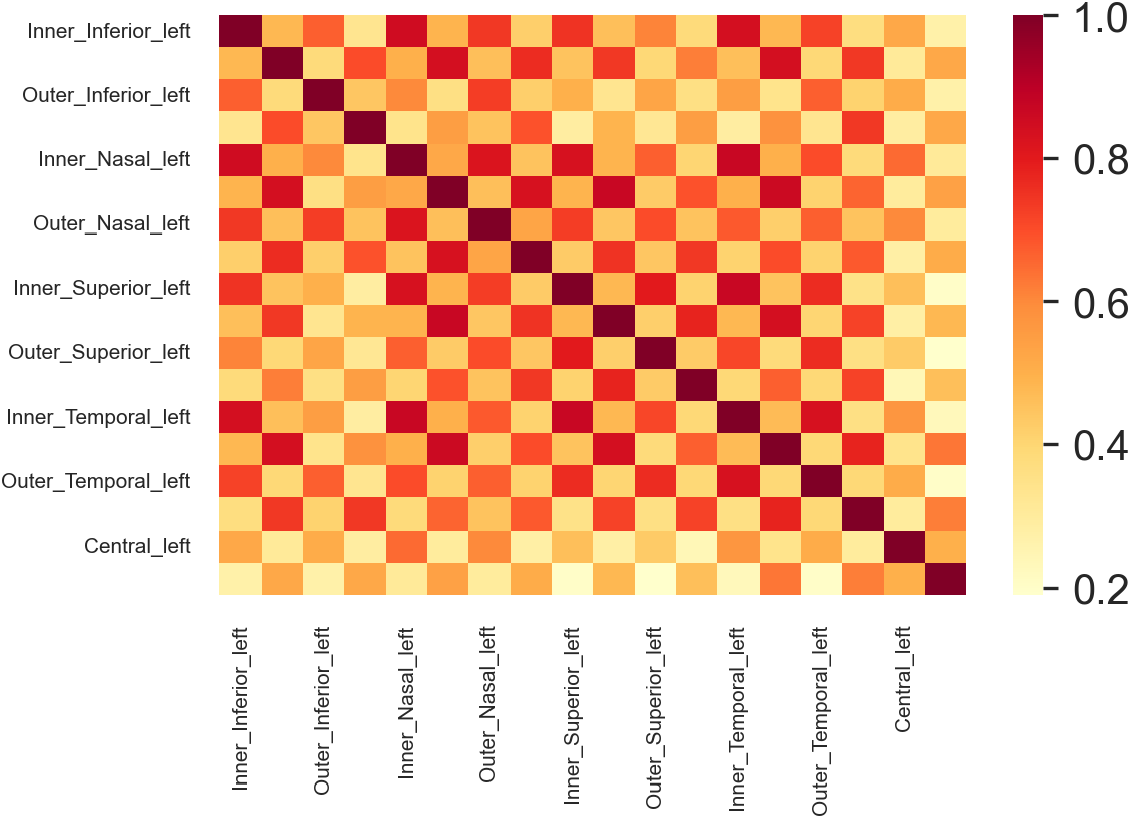
Covariance matrix of macular subfields

**Figure S2.**
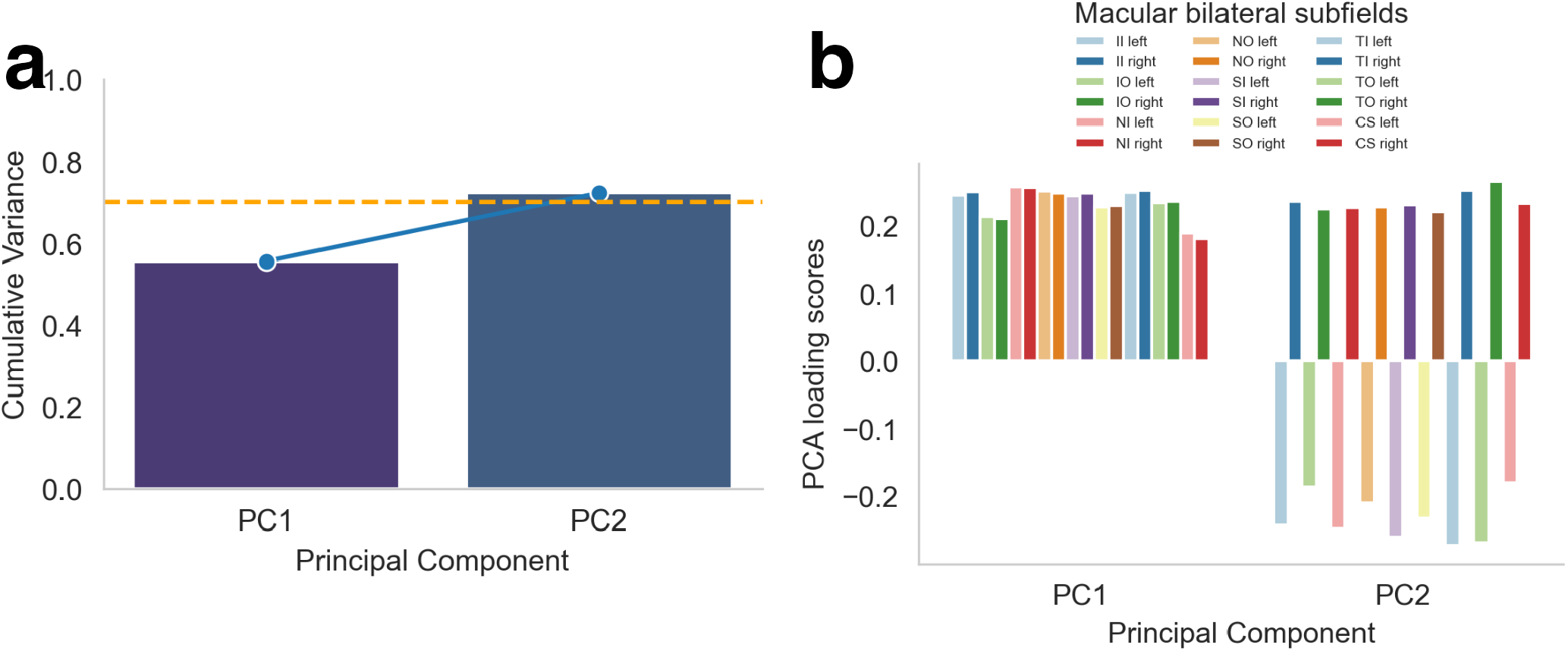
a. Cumulative percentage variance explained by the principal components. The first two and four principal components respectively account for more than 70 percent variance, as indicated by the horizontal dotted red line. These components were included in the subsequent regression models. b. The loading scores of macular subfields for the first two principal components for each eye respectively

**Figure S3.**
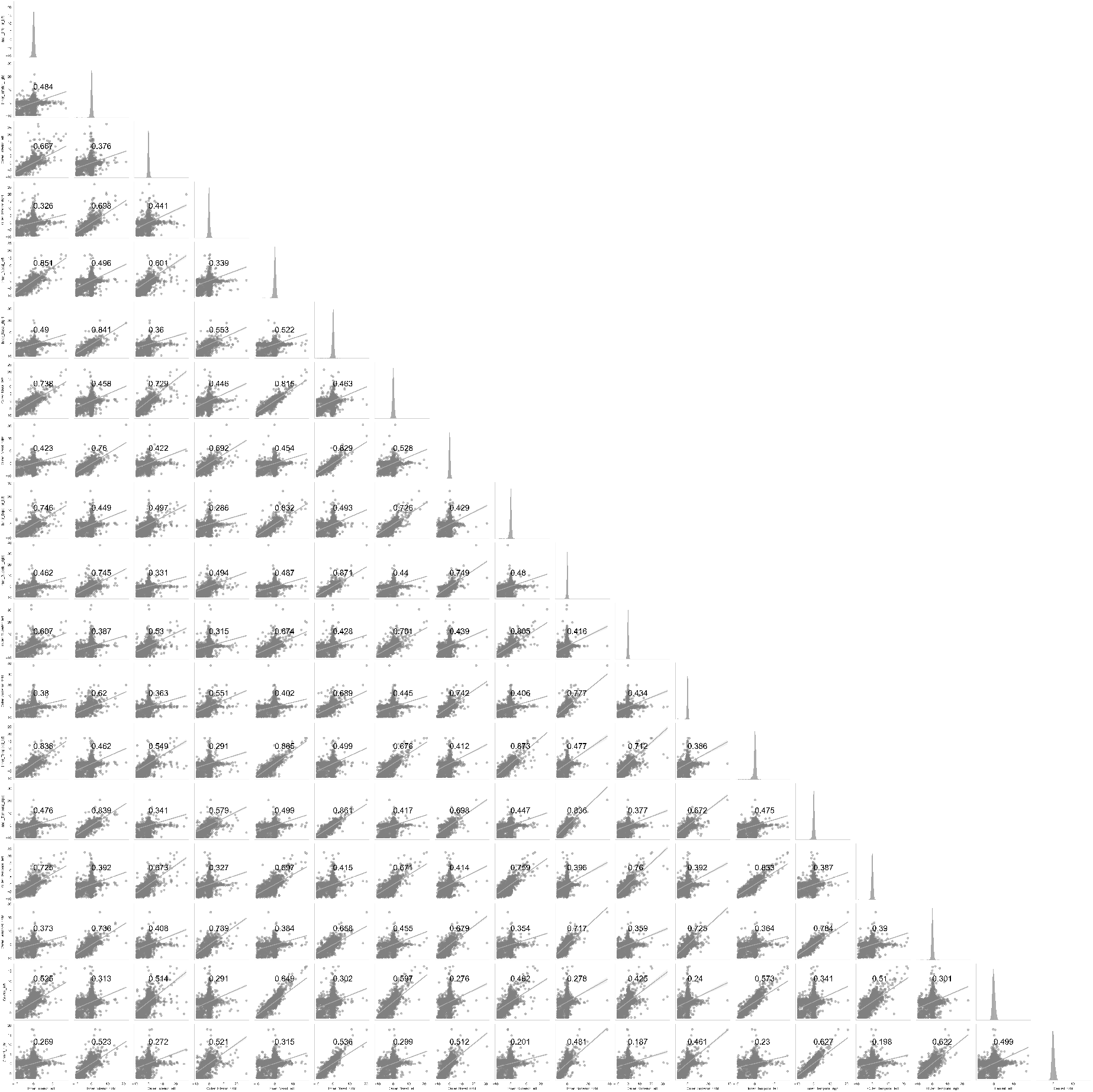
Pearson correlation coefficients between retinal phenotypes and showing their individual distribution

**Figure S4.**
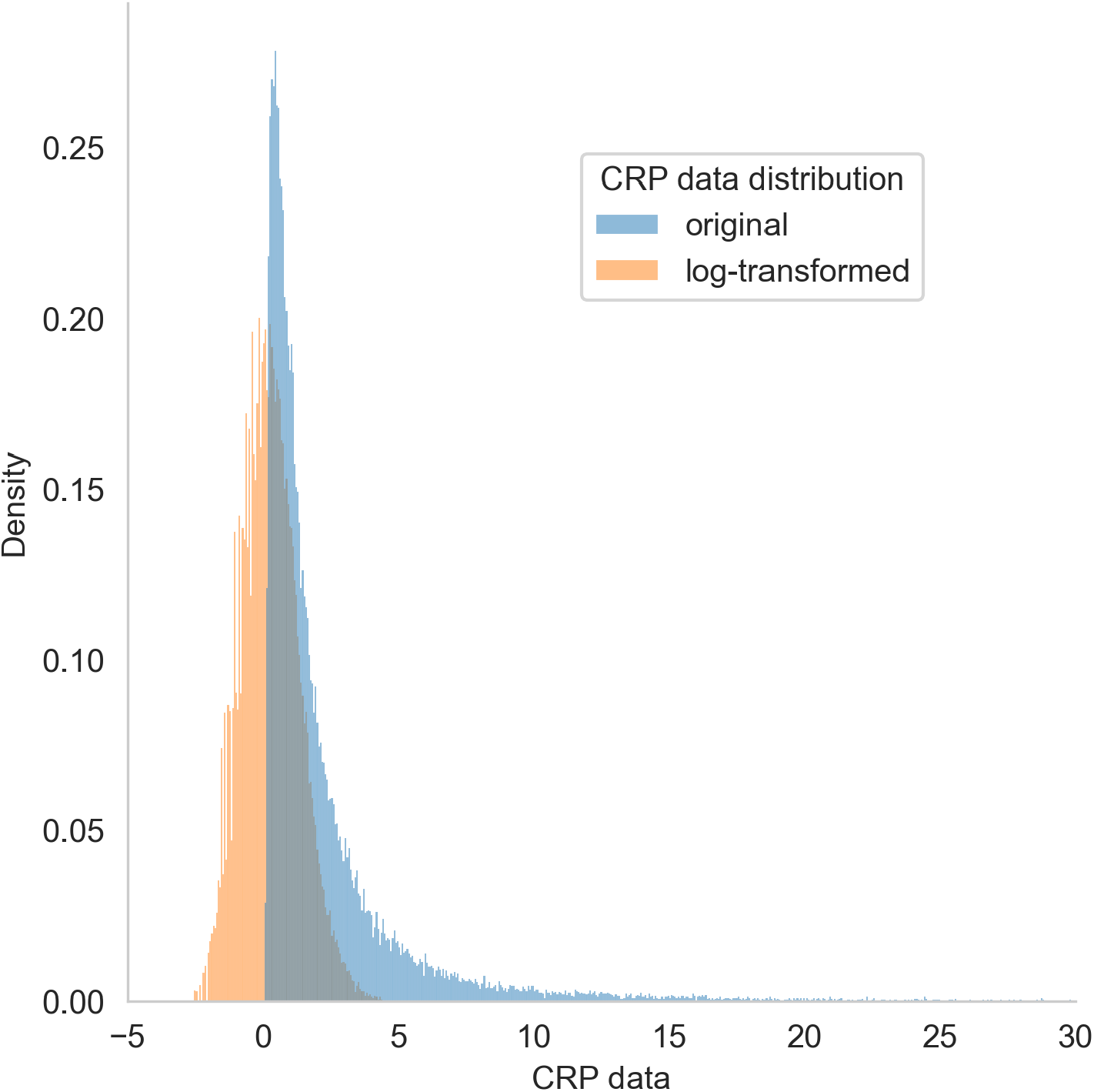
Logarithmic transformation of CRP levels

